# A Registry-Based Dataset of Hypertension and Type 2 Diabetes Patients from Selected Public Health Centres in Addis Ababa, Ethiopia

**DOI:** 10.64898/2026.09.02.26362084

**Authors:** Nathnael T Tuffa, Pavel Vazquez, Abdissa T Bedada, Simon Rayner

## Abstract

Hypertension and Type 2 Diabetes are two of the most prevalent non-communicable diseases in the world and leading causes of morbidity and mortality. Diabetes is the most common global metabolic disorder and is considered as a major public health threat. Although both conditions are becoming more prevalent in the sub-Saharan Africa, descriptive datasets with large number of entries are not readily available. We gathered data from a total of 13 public health centers located in eight of the eleven sub-cities of Addis Ababa, Ethiopia, and included those patients who were registered in a period from September 2021 to June 2022. From a total dataset of 3785 patients, 2242 had hypertension, 990 had diabetes, and 553 had both conditions. To our knowledge, no comparable Ethiopian hypertension and Type 2 diabetes registry dataset has previously been made publicly available; by releasing these data, we provide a resource for future hypertension, diabetes, multimorbidity, and population-health research.

|  |  |
| --- | --- |
| Design Type(s) | Retrospective |
| Measurement Type(s) | Blood Pressure, Fasting Blood Sugar, Height, Weight |
| Technology Type(s) | Sphygmomanometer, Glucometer |
| Factor Type(s) | Diagnosis |
| Sample Characteristic - Organism | Homo sapiens |
| Sample Characteristic - Location | Ethiopia |

## Background and Summary

Non-communicable diseases (NCDs) are currently responsible for roughly 74% of all deaths globally, representing a significant shift in global health over the last few decades[1]^1^. Of these, Hypertension (HTN) and Type 2 Diabetes (T2D) are two of the most prevalent[2]. The International Diabetes Federation (IDF) estimates that 537 million adults (20-79 years) are living with diabetes. This number is predicted to rise to 643 million by 2030 and 852.5 million by 2050[3]. At the same time, hypertension is an even more common problem. According to WHO, in 2019 the global estimate of hypertension was 1.28 billion for adults aged between 30 and 79[4].

However, what these numbers fail to capture is the disparity that exists between High Income Countries (HICs) and Low/Middle Income Countries (LMICs) both in terms of disease incidence and research capacity to study these diseases. For example, it is estimated that 75% of NCDs and ~86% of early NCD related deaths (i.e., before age 70) occur in LMICs^2^. A similar imbalance exists in research capacity, often referred to as the 10/90 gap (i.e., 10% of global research funding is spent on diseases that are responsible for 90% of the global health burden). Within the context of NCDs the difference is even more striking – recent WHO data indicates that only 0.2% of NCD funding is allocated to institutions in LMICs[5]. The problem is further compounded by the fact that most trials and studies are carried out in populations from HICs. This creates a “contextual evidence gap” where findings and derived treatments do not necessarily extend to other ancestries[6]. Finally, while many infectious diseases have been eradicated, or dramatically reduced in terms of impact in HICs, LMICs face the additional challenge of dealing with a double disease burden^6^ and interpreting the impact of comorbidity in NCDs, e.g.[7][8].

Thus, as both T2D and HTN are becoming more prevalent in sub-Saharan Africa, there is a paucity of open-access, patient-level datasets from this region. Moreover, while many studies are published each year on these topics from this region, it does not seem common practice to publish the complete data that formed the basis for the study. For instance, a 2020 Ethiopian study[9] reviewed and summarized the medical records from 3056 diabetic patients (both type 1 and type 2) but the raw data was not made available. This lack of accessible data exacerbates the contextual evidence gap, particularly given emerging evidence that key clinical and phenotypic baselines—such as the relationship between Body Mass Index (BMI) and T2D onset—vary significantly across ancestral populations and cannot be extrapolated from HIC cohorts[10], [11]

Furthermore, in the absence of centralized Electronic Health Records (EHR) in this setting, constructing usable cohorts requires intensive manual digitization from paper-based registries. In this Data Descriptor, we report and characterize a curated baseline registry dataset of T2D and HTN patients collected across 13 selected public health centers spanning 8 of the 11 sub-cities in Addis Ababa, Ethiopia. The primary objective of this paper is to describe the structure, completeness, and reusability of this dataset and provide a transparent phenotypic baseline for future epidemiological and genetic studies in underrepresented populations.

## Methods

The data in this study were collected from the “hypertension and diabetes” national registry of the Ministry of Health of Ethiopia. This is a paper-based registry that is present in all government health centers and records all baseline data of patients with T2D and HTN together with their follow up visits every three months. We used data from selected health centers in 8 sub-cities of Addis Ababa and included those patients who were registered in the period of September 2021 to June 2022. Patient identifying information was not included in the dataset, thus the data is completely anonymous.

### Data Collection and Ethical Approval

This study involved the retrospective extraction and digitization of routine primary care records. Data collection was conducted in accordance with institutional guidelines and governed by administrative permissions approved during study initiation. Because the study relied strictly on secondary, de-identified data with zero personally identifiable information (PII) extracted or stored, a waiver of explicit written consent was applicable under national ethical guidelines for retrospective chart reviews. This was obtained from the Medical Biochemistry Departmental Ethics Review Committee (DERC) and the institution review board (IRB) of College of Health Sciences Addis Ababa University (approval number CHS/RTTD/76/2022).

### Data Summary

The data presented in this study were collected in accredited government health centers and represent basic de-identified patient information. We gathered data from a total of 13 health centers located in eight of the eleven sub-cities in Addis Ababa, Ethiopia between September 2021 to June 2022. The health centres represent most of the city, but were primarily selected based on existing clinical relationships with the centres. The data was manually entered into a spreadsheet by physicians and the initial dataset consisted of information on 3965 patients (raw_registry_data.csv) and excluded any identifying information such as date of birth, address and ID number. This large and diverse dataset allowed us to gain a comprehensive understanding of the characteristics and health outcomes of patients with hypertension and diabetes in the region and aid in the prioritization of patients suitable for inclusion in a subsequent study to perform deep genetic profiling of T2D patients in Ethiopia. The dataset is available at doi.org/10.5281/zenodo.20542297[12]

### Data window

We collected data between September 2021 to June 2022.

### Selected health centers

The included health centers were *Kolfe Keranyo Woreda 03, Kolfe Keranyo Woreda 06, Kolfe Keranyo Woreda 04, Kolfe Keranyo Woreda 08, Kirkos Woreda 11, Lideta Woreda 01, Lideta Woreda 06, Lideta Woreda 10, Nifas silk-Laphto Woreda 02, Arada Woreda 04, Bole Woreda 12, Gulele Woreda 09*, and *Addis Ketema Woreda 08*.

### Inclusion and exclusion criteria

Our inclusion criteria were diagnosis of T2D and/or HTN and complete documentation of baseline data (Table 1). The diagnoses of both diseases were based on the WHO guideline by general practitioners working in the “chronic diseases’ clinics” in the selected health centers. T2D patients had 2 records of fasting blood sugar greater or equal to 126mg/dl and/or had a HbA1C level greater or equal to 6.5%. HTN patients were on anti-hypertensive medications and had systolic blood pressure records of greater or equal to 140mmHg and/or diastolic blood pressure records greater or equal to 90mmHg. BMI were calculated using the recorded weight and height. This study included only adult patients as these are the only ones treated in the chronic diseases’ clinics.

**Table 1.** Description of the columns of the dataset.

| Name | Type | Description |
| --- | --- | --- |
| ID | Numerical | Unique anonymized Identification Number. |
| Sex | Object | M for male and F for female. |
| Age | Numerical | Age in number of years. |
| Weight | Numerical | Weight in kilograms (kg). |
| Height | Numerical | Height in centimetres (cm). |
| BMI | Numerical | Body mass index calculated as:<br>$\text{Weight}[\text{kg}]/\text{Height}^2[\text{cm}^2]$ |
| T2D | Boolean | Type 2 Diabetes diagnosed. 1 for presence and 0 for absence. |
| HTN | Boolean | Hypertension diagnosed. 1 for presence and 0 for absence. |
| Area | Text | Area where the patient was admitted for consultation. Consisting of administrative division followed by postal code. |
| Precondition | Text | Free text for submission of other known preconditions. |

Our initial dataset comprised 4019 patient records. The data was filtered for duplicates (47 records), missing data (126 records), and patients who were diagnosed as being type 1 diabetes or prediabetic (7 records). This left us with a filtered dataset of 3839 patient records (Registry_Data_final.csv).

### Methodology

The data were previously recorded in the public paper-based registries as per the guidelines of the Ethiopian Ministry of Health that instruct the collection of baseline and follow up data of all registered diabetes and hypertension patients attending the clinics. The data were collected and entered into the registries by the treating general practitioners and nurses in the respective clinics. The weight and height of the patients were measured using calibrated digital weight scales with a built-in height measuring analogue meter. Blood pressure was measured by mercury sphygmomanometers, while blood sugar was measured in the respective laboratories using electronic blood glucose meters.

## Data records

### Data description

A description of our dataset parameters is presented in Table 1. No information allowing identification of patients was collected.

### Dataset Composition

We then used the Z-Score [13] to perform an outlier analysis on the ‘Age’, ‘Weight’ and ‘Height’ variables to identify any anomalous entries that could be attributed to potential input errors. We used a threshold of three standard deviations from the mean to flag any entries that deviated significantly from the norm. In total, we removed 63 entries, ensuring that our dataset was free from any significant outliers that could have distorted our results.

The final filtered dataset contained entries for 3776 patients, of which 2236 had HTN, 987 had T2D, and 553 had both conditions (Figure 1). Summary statistics are provided in Table 2.

**Table 2.**
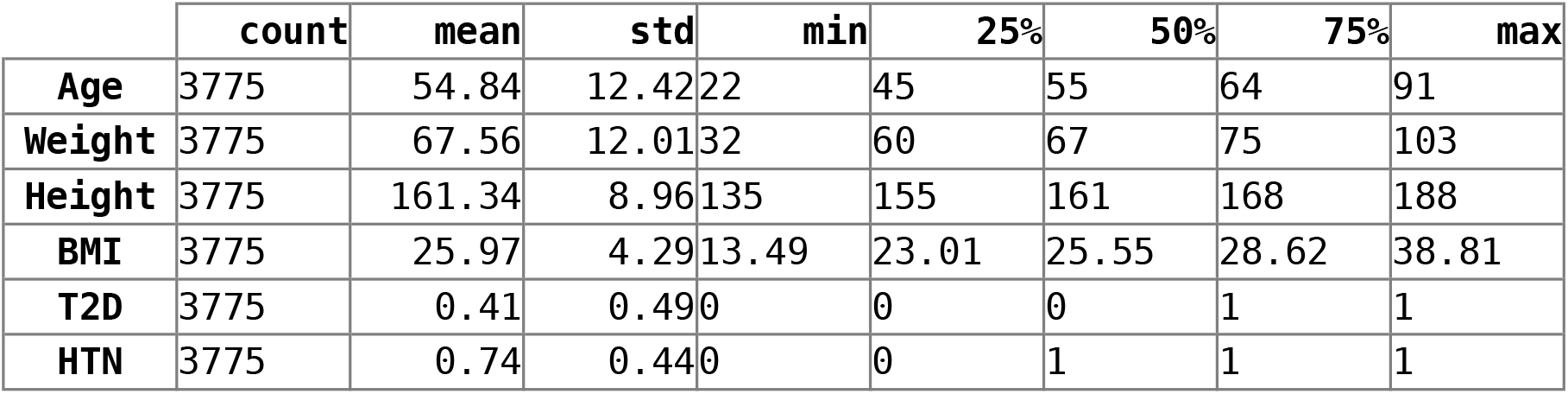
Summary statistics of dataset.

**Figure 1.**
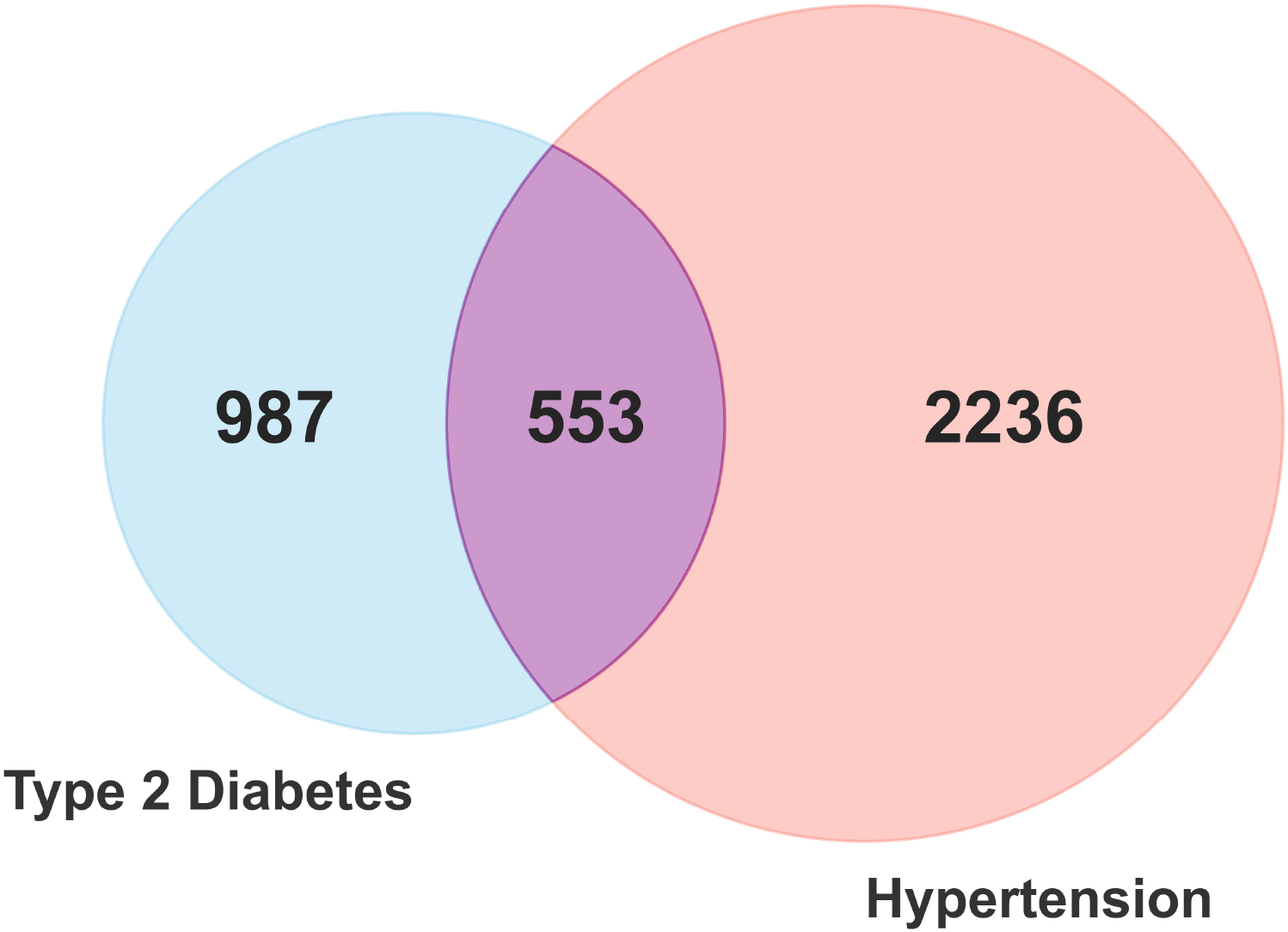
Proportion of patients with type 2 diabetes, hypertension and both conditions

## Technical validation

Data Normality, Feature Covariation, and Quality Control

To characterize the distributions and relationships among the main variables in the dataset (Age, Weight, Height, and BMI), pairplot matrix analyses were generated, stratified by hypertension diagnosis (Figure 2) and Type 2 Diabetes diagnosis (Figure 3).

**Figure 2.**
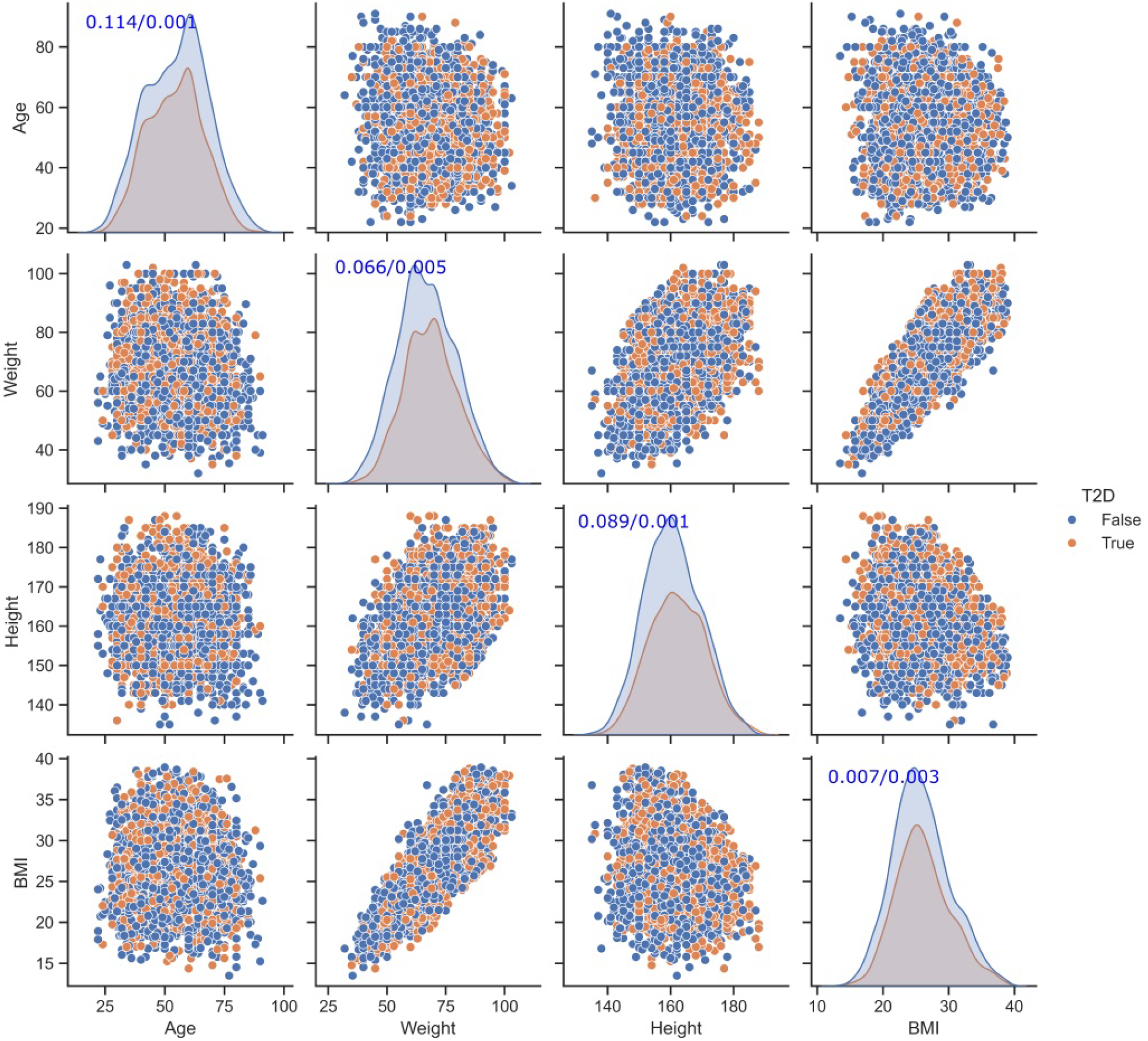
Data distribution among variables, based on hypertension diagnosis. The diagonal shows a univariate distribution plot to show the marginal distribution of the data in each column. Numbers in brackets along the diagonal show the results of the Lilliefors (adapted Kolmogorov-Smirnov) test for normality and the corresponding p-value

**Figure 3.**
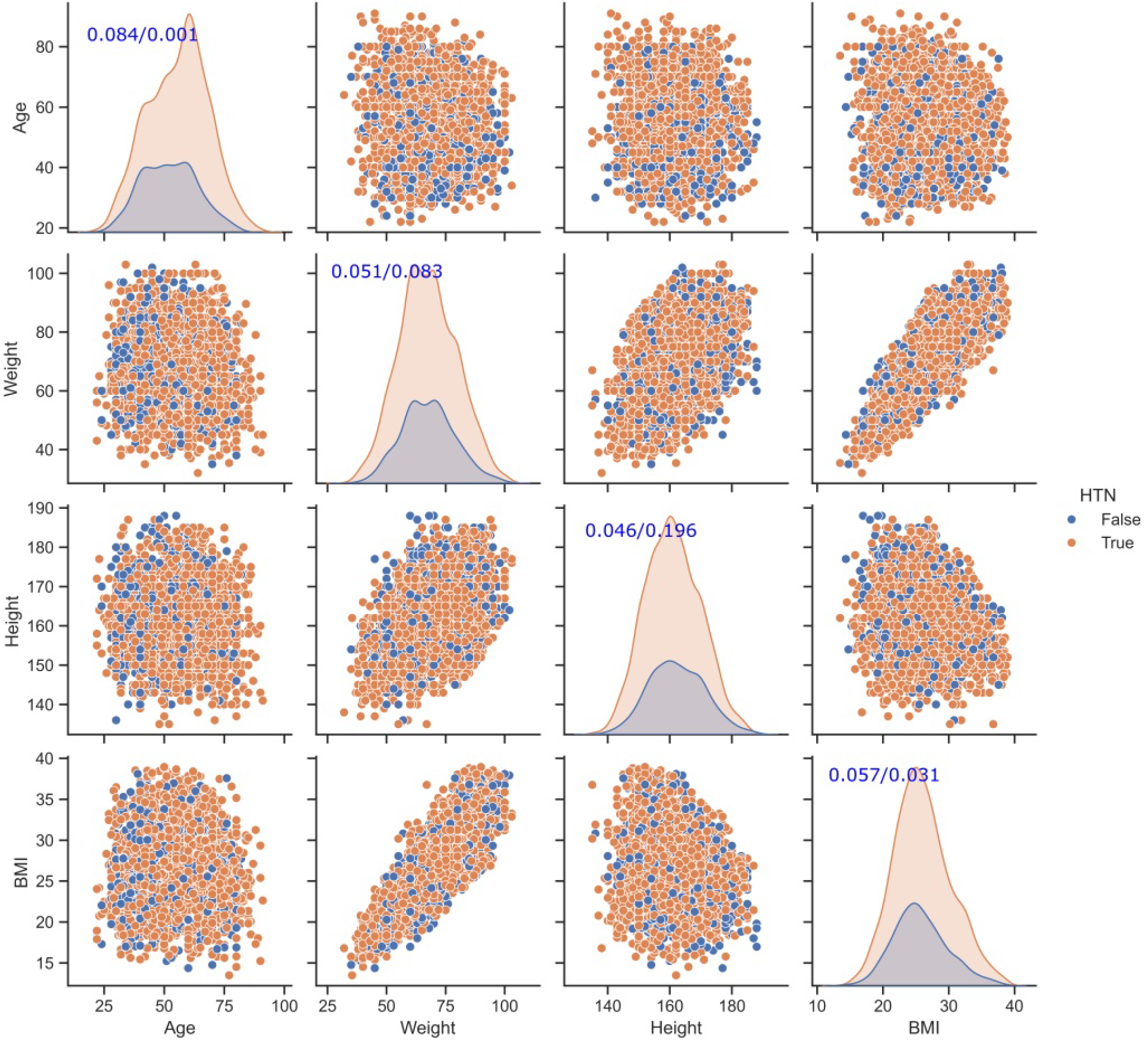
Data distribution among variables, based on type 2 diabetes diagnosis. The diagonal shows a univariate distribution plot to show the marginal distribution of the data in each column. Numbers in brackets along the diagonal show the results of the Lilliefors (adapted Kolmogorov-Smirnov) test for normality and the corresponding p-value

The diagonal univariate density plots display the marginal distributions for each physical variable. To guide future secondary data users on appropriate statistical modeling (i.e., selecting parametric vs. non-parametric tests), normality was formally evaluated along the diagonal using the Lilliefors test (adapted Kolmogorov-Smirnov). These tests are included simply to characterise the statistical properties of the dataset, rather than draw any clinical conclusions.

The off-diagonal scatter plots provide a visual overview of relationships among the recorded variables. In addition, these plots were used as part of the data-quality assessment process to identify implausible values and potential data-entry artefacts, such as swapped height and weight measurements. Visual inspection indicated that the cleaned dataset contained no obvious physiological inconsistencies and retained the expected relationships among height, weight, and BMI.

To compare our dataset to previously published studies, we searched PubMed for publications containing “type 2 diabetes” and “Ethiopia” in the Title/Abstract field. This returned a set of 105 hits. These were then reviewed manually to identify which studies reported an associated dataset. We found 14 such reports [14], [15], [16], [17], [18][19], [20][21], [22], [23], [24], [25], [26], of which the largest available dataset, [26], contained 879 participants. This is significantly less than the 3776 patients in our own dataset.

## Limitations

While this dataset provides a unique, digitized primary care registry across 8 sub-cities in Addis Ababa, prospective users should recognise the following caveats:

### Convenience sampling and representativeness

The health centres included in this study were selected based on practical considerations and existing clinical collaborations. Consequently, the dataset was not designed to estimate disease prevalence and should not be considered representative of Addis Ababa or Ethiopia as a whole. In particular, it may not reflect patterns of healthcare utilisation among patients attending private healthcare facilities or individuals not accessing healthcare services.

### Challenges associated with Paper-Based Records

Due to decentralized, paper-based logging in primary care facilities, certain longitudinal and behavioral variables (e.g., smoking history, detailed socioeconomic status, exact disease duration) were inconsistently recorded and excluded to maintain high data completeness.

### Extreme Value Filtering

Z-score filtering was applied to address clear transcription errors in paper charts; Thus, extreme clinical presentations may have been excluded.

### Cross-Sectional Baseline

This dataset represents a static baseline registry capture and does not currently track long-term longitudinal clinical outcomes or mortality.

Despite these limitations, the dataset provides a rare and openly available registry-derived resource describing hypertension and Type 2 diabetes patients attending public health centres in Addis Ababa. We believe it will be of value for future research into hypertension, diabetes, multimorbidity, and population health in Ethiopia.

## Data Availability

All data produced are available online at
The dataset is available at https://zenodo.org/records/20542297

## Code Availability

The code used in the analysis for this study is open-source and freely available at: https://github.com/CBGOUS/T2D-HTN-analysis

## Acknowledgements

We would like to acknowledge the contributions of Isa Salo Abdo, MD, and Megan Girma Gemechu, MD, from Addis Ababa University, as well as Yacob Alemu Bizuneh, MD, from Debre Markos University.

## Author Contributions

NTT and ATB collected data. PV, NTT and SR curated and analysed the data. EAA conceived the idea and checked the accuracy of the data. All authors contributed to the writing and revision of the manuscript.

## Competing Interests

The authors declare no competing interests.

## Notes

### Competing Interest Statement

The authors have declared no competing interest.

## References

[1] OECD, “The Health and Economic Benefits of Tackling Non-Communicable Diseases,” OECD Health Policy Studies, Apr. 2026, doi: 10.1787/e20cbbc3-en.

[2] J. Bai, J. Cui, F. Shi, and C. Yu, “Global Epidemiological Patterns in the Burden of Main Non-Communicable Diseases, 1990-2019: Relationships With Socio-Demographic Index,” Int J Public Health, vol. 68, p. 1605502, 2023, doi: 10.3389/ijph.2023.1605502.

[3] International Diabetes Federation, “IDF Diabetes Atlas, 11th edn.” Brussels, Belgium: International Diabetes Federation, 2025.

[4] B. Zhou et al., “Worldwide trends in hypertension prevalence and progress in treatment and control from 1990 to 2019: a pooled analysis of 1201 population-representative studies with 104 million participants,” The Lancet, vol. 398, no. 10304, pp. 957–980, Sep. 2021, doi: 10.1016/S0140-6736(21)01330-1.

[5] “Investments on grants for biomedical research by funder, type of grant, health category and recipient in 2020.” Accessed: May 09, 2026. [Online]. Available: https://www.who.int/observatories/global-observatory-on-health-research-and-development/monitoring/investments-on-grants-for-biomedical-research-by-funder-type-of-grant-health-category-and-recipient

[6] B. S. Hasan, M. A. Rasheed, A. Wahid, R. K. Kumar, and L. Zuhlke, “Generating Evidence From Contextual Clinical Research in Low-to Middle Income Countries: A Roadmap Based on Theory of Change,” Front. Pediatr., vol. 9, Dec. 2021, doi: 10.3389/fped.2021.764239.

[7] J. E. Akumu et al., “Diabetes mellitus, TB, and HIV multi-morbidities among adults in Uganda,” Public Health Action, vol. 16, no. 1, pp. 63–68, Mar. 2026, doi: 10.5588/pha.25.0032.

[8] U. P. Gujral et al., “Diabetes screening among people with tuberculosis: a systematic review and meta analysis,” eClinicalMedicine, vol. 93, Mar. 2026, doi: 10.1016/j.eclinm.2026.103803.

[9] G. Gebremedhin, F. Enqueselassie, N. Deyessa, and H. Yifter, “Urban-Rural Differences in the Trends of Type 1 and Type 2 Diabetes Among Adults Who Received Medical Treatment from Public Hospitals in Resource-Poor Community Tigray, Ethiopia,” DMSO, vol. Volume 13, pp. 859–868, Mar. 2020, doi: 10.2147/DMSO.S238275.

[10] G. Gebremedhin, F. Enqueselassie, N. Deyessa, and H. Yifter, “Urban-Rural Differences in the Trends of Type 1 and Type 2 Diabetes Among Adults Who Received Medical Treatment from Public Hospitals in Resource-Poor Community Tigray, Ethiopia,” DMSO, vol. Volume 13, pp. 859–868, Mar. 2020, doi: 10.2147/DMSO.S238275.

[11] B. Orazumbekova et al., “Evidence of ethnic variations in the relationships between routinely recorded clinical factors and T2D: a systematic review and meta-analysis,” Int J Obes, vol. 49, no. 10, pp. 1929–1945, Oct. 2025, doi: 10.1038/s41366-025-01848-9.

[12] P. V. Faci, N. T. Tuffa, and S. Rayner, “Hypertension and Diabetes Registry Dataset in Addis Ababa, Ethiopia”, doi: 10.5281/zenodo.20542297.

[13] A. E. Curtis, T. A. Smith, B. A. Ziganshin, and J. A. Elefteriades, “The Mystery of the Z-Score,” Aorta (Stamford), vol. 4, no. 4, pp. 124–130, Aug. 2016, doi: 10.12945/j.aorta.2016.16.014.

[14] T. G. Haile, T. Mariye, D. B. Tadesse, G. G. Gebremeskel, G. G. Asefa, and T. Getachew, “Prevalence of hypertension among type 2 diabetes mellitus patients in Ethiopia: a systematic review and meta-analysis,” International Health, vol. 15, no. 3, pp. 235–241, May 2023, doi: 10.1093/inthealth/ihac060.

[15] A. S. Tegegne, “Joint Predictors of Hypertension and Type 2 Diabetes Among Adults Under Treatment in Amhara Region (North-Western Ethiopia),” Diabetes Metab Syndr Obes, vol. 14, pp. 2453–2463, 2021, doi: 10.2147/DMSO.S309925.

[16] M. S. Erkocho, D. T. Adugna, T. T. Arficho, and A. G. Azene, “Poor dietary practice and associated factors among type-2 diabetes mellitus patients on follow-up in Nigist Eleni Mohammed Memorial Teaching Hospital, Ethiopia,” Pan Afr Med J, vol. 41, p. 164, 2022, doi: 10.11604/pamj.2022.41.164.28675.

[17] T. Bizuayehu, T. Menjetta, and M. Mohammed, “Obesity among type 2 diabetes mellitus at Sidama Region, Southern Ethiopia,” PLOS ONE, vol. 17, no. 4, p. e0266716, Apr. 2022, doi: 10.1371/journal.pone.0266716.

[18] A. K. Sendekie, A. K. Netere, and E. A. Belachew, “Hypoglycemic events and glycemic control effects between NPH and premixed insulin in patients with type 2 diabetes mellitus: A real-world experience at a comprehensive specialized hospital in Ethiopia,” PLoS One, vol. 17, no. 9, p. e0275032, 2022, doi: 10.1371/journal.pone.0275032.

[19] S. A. Kebede, B. S. Tusa, A. B. Weldesenbet, Z. T. Tessema, and T. A. Ayele, “Incidence of Diabetic Nephropathy and Its Predictors among Type 2 Diabetes Mellitus Patients at University of Gondar Comprehensive Specialized Hospital, Northwest Ethiopia,” J Nutr Metab, vol. 2021, p. 6757916, 2021, doi: 10.1155/2021/6757916.

[20] Y. Akalu and Y. Belsti, “Hypertension and Its Associated Factors Among Type 2 Diabetes Mellitus Patients at Debre Tabor General Hospital, Northwest Ethiopia,” Diabetes Metab Syndr Obes, vol. 13, pp. 1621–1631, 2020, doi: 10.2147/DMSO.S254537.

[21] K. G. Kiros, G. Y. Abyu, D. S. Belay, M. H. Goyteom, and T. K. Welegebriel, “Magnitude of overweight and associated factors among type 2 diabetes mellitus patients at Mekelle public hospitals, Tigray, Ethiopia: a cross-sectional study,” BMC Res Notes, vol. 12, no. 1, p. 762, Nov. 2019, doi: 10.1186/s13104-019-4791-1.

[22] G. G. Gebremeskel et al., “Magnitude of metabolic syndrome and its associated factors among patients with type 2 diabetes mellitus in Ayder Comprehensive Specialized Hospital, Tigray, Ethiopia: a cross sectional study,” BMC Res Notes, vol. 12, p. 603, Sep. 2019, doi: 10.1186/s13104-019-4609-1.

[23] S. Letta, F. Aga, T. A. Yadeta, B. Geda, and Y. Dessie, “Correlates of Glycemic Control Among Patients With Type 2 Diabetes in Eastern Ethiopia: A Hospital-Based Cross-Sectional Study,” Front. Endocrinol., vol. 13, Jul. 2022, doi: 10.3389/fendo.2022.939804.

[24] S. Hailemariam, T. Melak, and M. Abebe, “Waist Circumference Cutoff Point Determination for Defining Metabolic Syndrome in Type 2 Diabetes Mellitus in Ethiopia,” EJIFCC, vol. 30, no. 1, pp. 48–58, Mar. 2019.

[25] B. Dagnew and Y. Yeshaw, “Predictors of isolated systolic hypertension among type 2 diabetes mellitus patients in Jimma University Specialized Hospital, Southwest Ethiopia,” BMC Res Notes, vol. 12, no. 1, p. 510, Aug. 2019, doi: 10.1186/s13104-019-4550-3.

[26] S. Letta, F. Aga, T. Assebe Yadeta, B. Geda, and Y. Dessie, “Self-care practices and correlates among patients with type 2 diabetes in Eastern Ethiopia: A hospital-based cross-sectional study,” SAGE Open Medicine, vol. 10, p. 20503121221107337, Jan. 2022, doi: 10.1177/20503121221107337.

